# Standardized Multicenter Critical Care Database Integrating Minute-Level Vital Signs, Laboratory Tests, Interventions, and Outcomes: Profile of the OneICU Database

**DOI:** 10.1101/2025.09.12.25335671

**Authors:** Takahiro Kinoshita, Yutaka Umemura, Makoto Watanabe, Kenichiro Uchida, Yuji Nishimoto, Shinshu Katayama, Yoshiaki Inoue, Hiroshi Kurosawa, Yutaka Igarashi, Akira Kodate, Keita Iyama, Shigeaki Inoue, Hirotada Kobayashi, Yasushi Nakamori

**Author notes:** **Corresponding author contact information:** Takahiro Kinoshita, MD, MPH MeDiCU, Inc., 1-15-23, Higashinakamoto, Higashinari-ku, Osaka 537-0021, Japan.

## Abstract

**Introduction:** Standrdized intensive care unit (ICU) databases with high frequency data collection from multicenter electronic medical records remain scarce. We aimed to describe the profile of newly developed OneICU database, which includes minute-level recordings of vital signs, laboratory values, interventions, and diagnosis codes, and to evaluate the importance of vital sign measurement frequency and laboratory data completeness for developing machine learning–based clinical decision support.

**Methods:** This retrospective, multicenter observational study collected critically ill patient data from 12 tertiary care hospitals from 2013 to 2025. Patient demographics, measurement frequency of vital signs and laboratory tests, as well as the availability of the Sequential Organ Failure Assessment (SOFA) score components were compared across three large ICU databases: OneICU, Medical Information Mart for Intensive Care (MIMIC)-IV and eICU. We then evaluated the prediction accuracy of five machine learning models to forecast hypotensive events 60–120 minutes in advance, defined as a median invasive mean arterial pressure (MAP) < 65 mmHg or vasopressor initiation, using different MAP sampling frequencies: OneICU 1 minute, OneICU 5 minute, OneICU hourly, MIMIC IV hourly, and eICU 5 minute.

**Results:** OneICU currently includes 152,269 ICU stays from 127,757 unique patients. Compared with MIMIC-IV and eICU, OneICU captured more frequent vital signs (minute-level vs. hourly in MIMIC-IV and every five minutes in eICU) and provided broader availability of SOFA score components. In particular, the respiratory component was available for 73.6 % of stays in OneICU versus 37.8 % in MIMIC IV and 30.9 % in eICU, and the liver component for 93.3 % versus 45.2 % and 41.3 %, respectively. The test-set area under the receiver operating characteristic curve was highest for the OneICU 1-minute model (0.942), followed by OneICU 5-minute model (0.939), OneICU hourly model (0.901), eICU 5-minute model (0.899), and MIMIC IV hourly model (0.799).

**Conclusions:** A high resolution, multicenter ICU database integrating minute level vital sign recordings with comprehensive SOFA score coverage is feasible and was associated with superior hypotension prediction performance. OneICU enables detailed analyses of ICU trajectories and addresses the current scarcity of large scale ICU data from Asian populations.

## Background

The use of comprehensive, high-resolution time-series data from electronic medical records (EMRs) has become integral to advancing the management of critically ill patients in the intensive care unit (ICU). Continuous streams of vital signs, laboratory results, and interventions recorded at minute-level intervals provide opportunities for precise assessment of disease severity, early detection of clinical deterioration, and personalized therapeutic strategies. Recent studies suggest that large-scale time-series databases can facilitate data-driven approaches and artificial intelligence (AI) models, leading to improvements in ICU care and patient outcomes (1–5).

Despite these advantages, the development of large scale time-series ICU databases remains challenging due to the heterogeneity of EMR systems, the technical complexity of data standardization, and privacy considerations. Prominent databases such as the Medical Information Mart for Intensive Care (MIMIC-IV) (6), the eICU Collaborative Research Database (eICU-CRD) (7), Amsterdam University Medical Center database (AmsterdamUMCdb) (8), and High time-resolution intensive care unit dataset (HiRID) (3) have fueled numerous studies; however, vital-sign sampling frequencies differ markedly, ranging from every 2 min in HiRID to approximately hourly in MIMIC IV, complicating direct comparisons of datasets and the models developed from them. Moreover, most available datasets originate from single institutions in Western countries, limiting their generalizability. Asian ICU databases remain scarce, and initiatives that integrate high resolution data from geographically diverse hospitals are virtually nonexistent. This lack of representative data from Asian populations can lead to gaps in evidence-based practice and preclude the development of fair and context-appropriate AI.

To address these issues, we developed OneICU, a large, multicenter ICU database that aggregates data from various Japanese tertiary care hospitals. In this first report, we outline the database design, data processing workflow, and patient characteristics, and present machine learning models for hypotension prediction, evaluating prediction accuracy against MIMIC IV and eICU.

## Methods

### Database development

OneICU is a large-scale, continuously evolving ICU database established within a separate multicenter retrospective study whose primary objective is to develop a machine-learning model to predict sudden deterioration in acute care settings. The present manuscript profiles the database—its design, contents, and data quality—and report early modeling works; the results of the final model using the full dataset will be published separately upon the study completion. As of December 2025, 37 tertiary care hospitals in Japan were participating in the study, with 12 having completed data collection and cleaning. The protocol for the original study was approved by the Kansai Medical University Ethics Committee (reference number: 2024015). Given the inclusion of historical admissions, the Institutional Review Board (IRB) approved reliance on institutional public notification, rather than direct re contact, contingent upon the posting of prominent opt out notices at each site. Each participating institution displayed an opt out notice to allow patients or their families to decline participation. All sites had submitted their opt out notices to their institutional IRBs before the first data transfer, and any records flagged by opt out were excluded from the analytic database before anonymization.

Patients were eligible if they were admitted to medical, surgical, emergency, pediatric, neonatal ICUs, or coronary care, stroke care, or high care units and had at least one vital sign measurement. Data collection was defined to begin on April 1, 2010, or on the date each ICU-specific EMR system was implemented, whichever was later. Accordingly, for the currently participating hospitals, 2013 is the earliest year of data contribution. The IRB also approved the secondary use of de-identified data.

### Data Standardization Workflow

A key challenge for developing the OneICU database was standardizing the substantial volume of data generated by heterogeneous EMR systems into a single schema of tables. We therefore adopted an extract–load–transform (ELT) paradigm rather than the traditional extract–transform–load pipeline (9). In the ELT design, de identified yet still unstructured data are first loaded in bulk into a secure cloud environment, and only then are they transformed to the target schema.

The data are categorized into three types: static, point time-series, and interval time-series variables. Static variables are those without repeated measurements, including age, sex, and diagnosis active upon ICU discharge. Point time-series variables capture repeated observations at distinct time points, such as vital signs and laboratory test results. Interval time-series variables require both a start and end time to reflect real-world clinical practice, such as infusion and ventilator settings. This strategy facilitated high-volume data processing and minimized local computational bottlenecks. Detailed procedures for each step (extraction, loading, and transformation) are provided in the Supplementary Methods.

### Relational Structure

The OneICU database comprises four tiers—patients, hospitals, hospital_admissions, and icu_stays (Figure 1). Patient level attributes are stored in patients (key: *subject_id*) and hospital characteristics are stored in hospitals (key: *hospital_id*). Each hospitalization is represented in hospital_admissions and linked to patients and hospitals via *subject_id* and *hospital_id*. Admission level diagnoses are stored in hospital_admission_diagnoses and reference hospital_admissions via *hospital_admission_id*. Each ICU encounter is represented in icu_stays (key: *icu_stay_id*), which serves as the parent table for ICU diagnoses (icu_diagnoses), physiological measurements (e.g., vital_measurements, body_weight_measurements), therapeutic interventions (e.g., infusions, mechanical_ventilations), and laboratory and clinical assessments (e.g., laboratory_tests_blood, gcs); these tables join to icu_stays by *icu_stay_id*.

**Figure 1.**
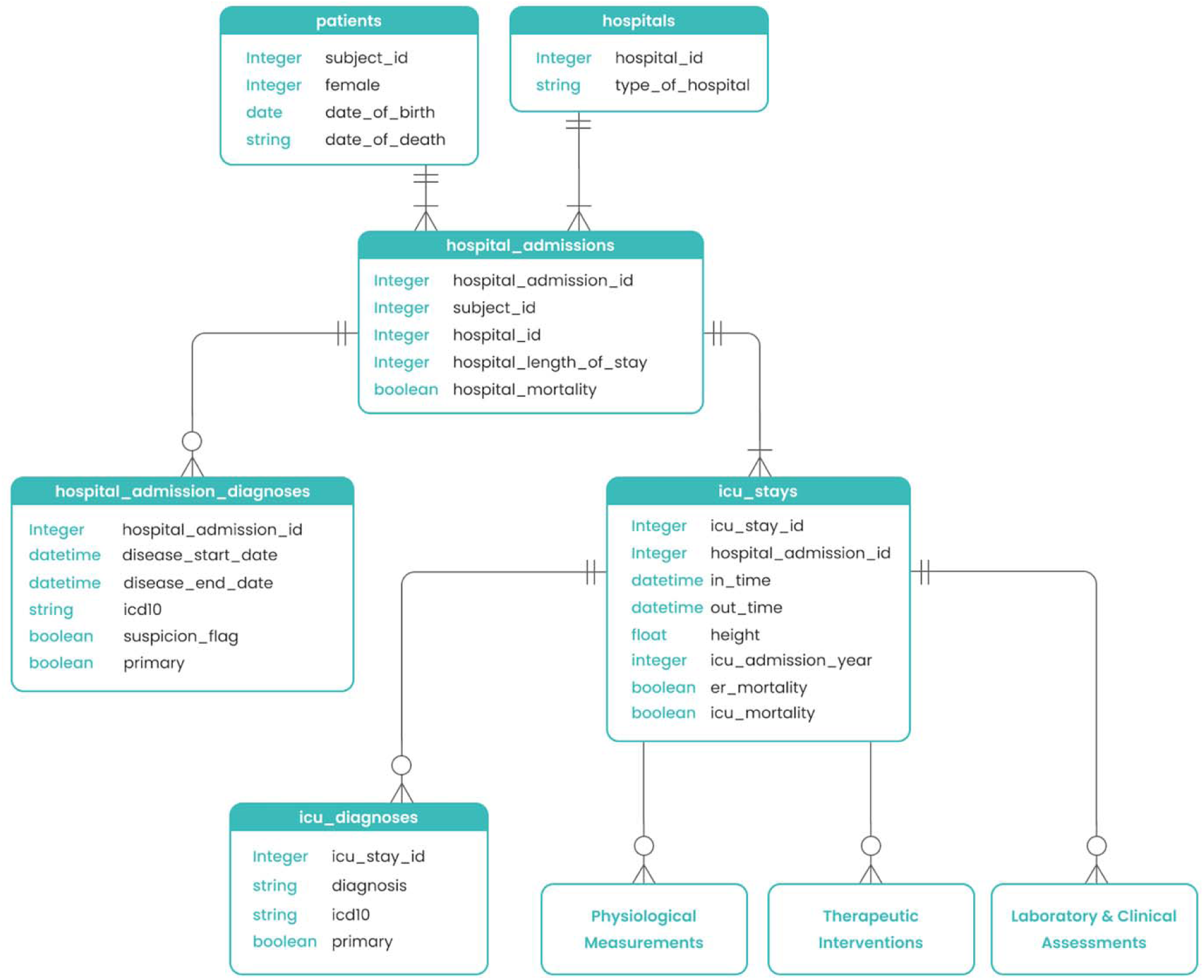
Entity–relationship diagram of the OneICU database Rectangles represent tables; listed fields are example column names and data types. One patient (*subject_id*) can have multiple hospital admissions (*hospital_admission_id*), and each admission can include multiple ICU stays (*icu_stay_id*). Each admission is associated with a single hospital (*hospital_id*). The icu_stays table anchors ICU diagnoses table and time stamped child tables for physiological measurements, therapeutic interventions, and laboratory and clinical assessments, all linked by *icu_stay_id*. Abbreviations: ICU, Intensive Care Unit

Descriptions of all variables across the tables are provided in the Supplementary Methods. We also generated 24 derived tables, including those providing the Charlson Comorbidity Index and hourly Sequential Organ Failure Assessment (SOFA) scores. The SQL code is publicly available in the project GitHub repository (https://github.com/medicu-inc/one-icu).

### Illustrative Example of Time-Series Data

To illustrate the granularity of OneICU data, we plotted vital signs, medication/infusion intervals, and severity scores (SOFA, Japanese Association for Acute Medicine Disseminated

Intravascular Coagulation [JAAM DIC]), which were calculated for all patients on cloud environment, for an arbitrary patient admitted with sepsis. Minute-level vital signs (heart rate [HR], blood pressure [BP], respiratory rate [RR], oxygen saturation [SpO ], body temperature [BT]) were extracted from the vital measurements table, while medications, fluids, and ventilator settings were accessed from interval time-series tables. Severity scores were plotted at 6-hour intervals to reflect clinical changes.

### Profile analysis

We compared the demographics of the OneICU database with two globally recognized and extensively studied benchmark ICU databases, MIMIC-IV and eICU. Comparisons included the number of centers, number of unique ICU stays, number of patients, data availability periods, patient age, sex, and ICU length of stays. We also assessed the availability and summary statistics of the SOFA score and the Acute Physiology and Chronic Health Evaluation (APACHE) II, III, and IVa scores. We quantified use of vasopressors (norepinephrine, epinephrine, vasopressin, dopamine, phenylephrine), invasive mechanical ventilation, noninvasive positive pressure ventilation (NPPV), high flow oxygen therapy, and continuous renal replacement therapy (CRRT). Among norepinephrine users, we compared the initial dose standardized to μg/kg/min. Mortality rates in the emergency room (ER), ICU, and in-hospital were investigated respectively. In some participating hospitals, patient records at ER of the severest cases were managed in the ICU-EMR system; therefore, we excluded patients who died in the ER from ICU and in-hospital mortality calculations.

The comprehensiveness of OneICU was evaluated by comparing the availability and frequency of key variables against MIMIC-IV and eICU. Specifically, we investigated the granularity of vital sign measurements by calculating the average measurements per hour for HR, RR, invasive and non-invasive BP, SpO , and BT among patients who stayed more than 24 hours. For laboratory tests, we calculated the average number of daily measurements of blood gas (pH, lactate), complete blood count (white blood cell count [WBC], hemoglobin [Hb]), chemistry (sodium, albumin), and coagulation (prothrombin time international normalized ratio, D-dimer). Availability of SOFA score components, respiratory, liver, cardiovascular, coagulation, renal, and central nervous system parameters, was also investigated.

### Machine learning

To assess the impact of vital-sign granularity on prediction model performance, we conducted comparative modeling across OneICU, MIMIC IV, and eICU to predict hypotensive events occurring 1–2 hours after the current observation window. Eligible ICU stays required recorded age and sex and at least one measurement of HR, RR, invasive MAP, and SpO . Each ICU stay was partitioned into consecutive, non overlapping 60 minute windows. For each window, we computed median HR, RR, and SpO and extracted the most recent available blood gas results (pH, partial pressure of oxygen [PaO ], partial pressure of carbon dioxide [PaCO ], base excess, lactate, sodium, potassium).

From OneICU, we extracted invasive MAP at three sampling frequencies: 60 values at 1 minute intervals, 12 values at 5 minute intervals, and 1 value at hourly intervals. In addition, we appended database specific MAP sequences for the other two databases: 1 value at hourly intervals (MIMIC IV) and 12 values at 5 minute intervals (eICU). The outcome was a hypotensive event occurring in the window 60–120 minutes after the index window, defined as either (i) median invasive MAP < 65 mmHg or (ii) initiation of a vasopressor. We excluded windows with missing invasive MAP, windows for which the outcome occurring 60–120 minutes after the index window could not be defined or ascertained, and all windows at or after the first vasopressor initiation to avoid treatment related label leakage; this was particularly required because eICU lacks infusion end times.

Within each database, data were randomly split into a training set (80%) and a test set (20%) at the ICU-stay level. In the training partition, we used H2O AutoML (H2O.ai, USA) to fit multiple algorithms with automated hyperparameter tuning (generalized linear models, gradient boosting machines, distributed random forests, extremely randomized trees, deep neural networks, and stacked ensembles), eliminating investigator-driven hyperparameter selection. To maintain computational efficiency, cross-validation was disabled and the training partition was further divided into mutually exclusive subsets for base training (80%), blending (10%), and the leaderboard (10%). Stacked ensembles were trained using the blending subset, and all candidate models were ranked by the area under the receiver operating characteristic curve (AUROC) computed on the reserved leaderboard subset. The final model was defined as the top ranked algorithm on this leaderboard, for the combination of database and sample frequency. For reporting, we retained a logistic regression baseline and the best performing machine-learning model. Performance on the held out test set was summarized by ROC curves with AUROC. We also evaluated specificity, positive predictive value (PPV), and negative predictive value (NPV) at the threshold corresponding to 90% sensitivity, and sensitivity, PPV, and NPV at the threshold corresponding to 90% specificity. All analytic steps described above were rigorously standardized across databases; only the sampling frequency of invasive MAP features differed.

Continuous variables are presented as medians (25–75 percentiles), and categorical variables as numbers (%). All analyses were performed using Google BigQuery (Google LLC, USA), R 4.3.1 (Institute for Statistics and Mathematics, Austria), and Python 3.12.3 (Python Software Foundation, USA). All code can be accessed through the following GitHub repository: https://github.com/takapion/OneICU_profile_paper.

## Results

Figure 2 displays the progression of vital signs, interventions, and severity scores during the ICU stay of a patient with septic shock caused by acute obstructive suppurative cholangitis. HR, systolic and diastolic BP, RR, and SpO_2_ fluctuated during the first 90 minutes, reflecting clinical instability and subsequent response to therapy. Norepinephrine was started to stabilize hemodynamics, followed by vasopressin for additional support, while mechanical ventilation was implemented based on the patient’s respiratory status. SOFA score and JAAM DIC score were elevated at ICU admission and both scores declined over time after antibiotic and anticoagulation medications.

**Figure 2.**
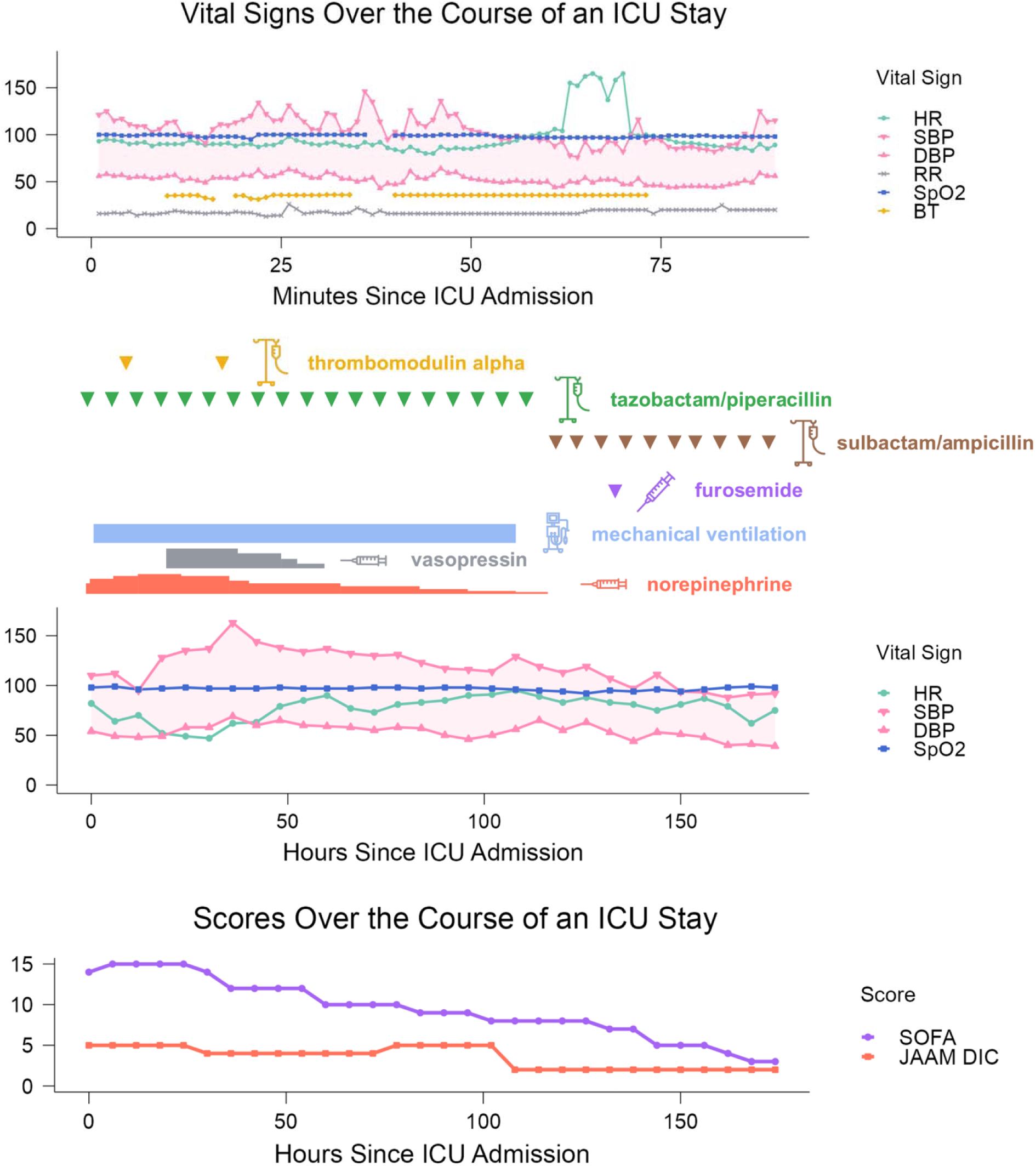
Example Patient’s ICU Stay: Vital Signs, Interventions, and Severity Scores The figure illustrates the clinical trajectory of a patient admitted to the ICU with acute obstructive suppurative cholangitis and septic shock. A minute-by-minute plot of vital signs captures dynamic changes in the patient’s clinical condition. The time-series data show fluctuations in vital signs, reflecting clinical instability and response to interventions. Vasopressor administration (noradrenaline and vasopressin), as well as the initiation and termination of mechanical ventilation, are indicated. Severity scores (SOFA and JAAM DIC) are plotted at 6-hour intervals, demonstrating a gradual improvement following treatment initiation. Abbreviations: ICU, Intensive Care Unit; HR, Heart Rate; SBP, Systolic Blood Pressure; DBP, Diastolic Blood Pressure; RR, Respiratory Rate; SpO , Oxygen Saturation; BT, Body Temperature; SOFA, Sequential Organ Failure Assessment; JAAM DIC, Japanese Association for Acute Medicine Disseminated Intravascular Coagulation

Table 1 compares key characteristics of the OneICU, MIMIC-IV, and eICU. Both OneICU and eICU collected data from multiple institutions, whereas MIMIC-IV drew from a single center. Among the three databases, eICU contained the largest number of ICU stays and unique patients, followed by OneICU, with MIMIC-IV having the smallest sample size. Patient severity, as measured by the Acute Physiology Score III, could be compared between OneICU and MIMIC-IV and appeared higher in OneICU. The proportions receiving vasopressors, invasive mechanical ventilation, and CRRT were higher in OneICU and MIMIC-IV than in eICU; NPPV and high-flow oxygen therapy were not evaluable in eICU. ICU and in-hospital mortality rates were highest in MIMIC-IV compared to the other two databases. The median ICU length of stay was slightly longer in MIMIC-IV than in OneICU and eICU.

**Table 1.**
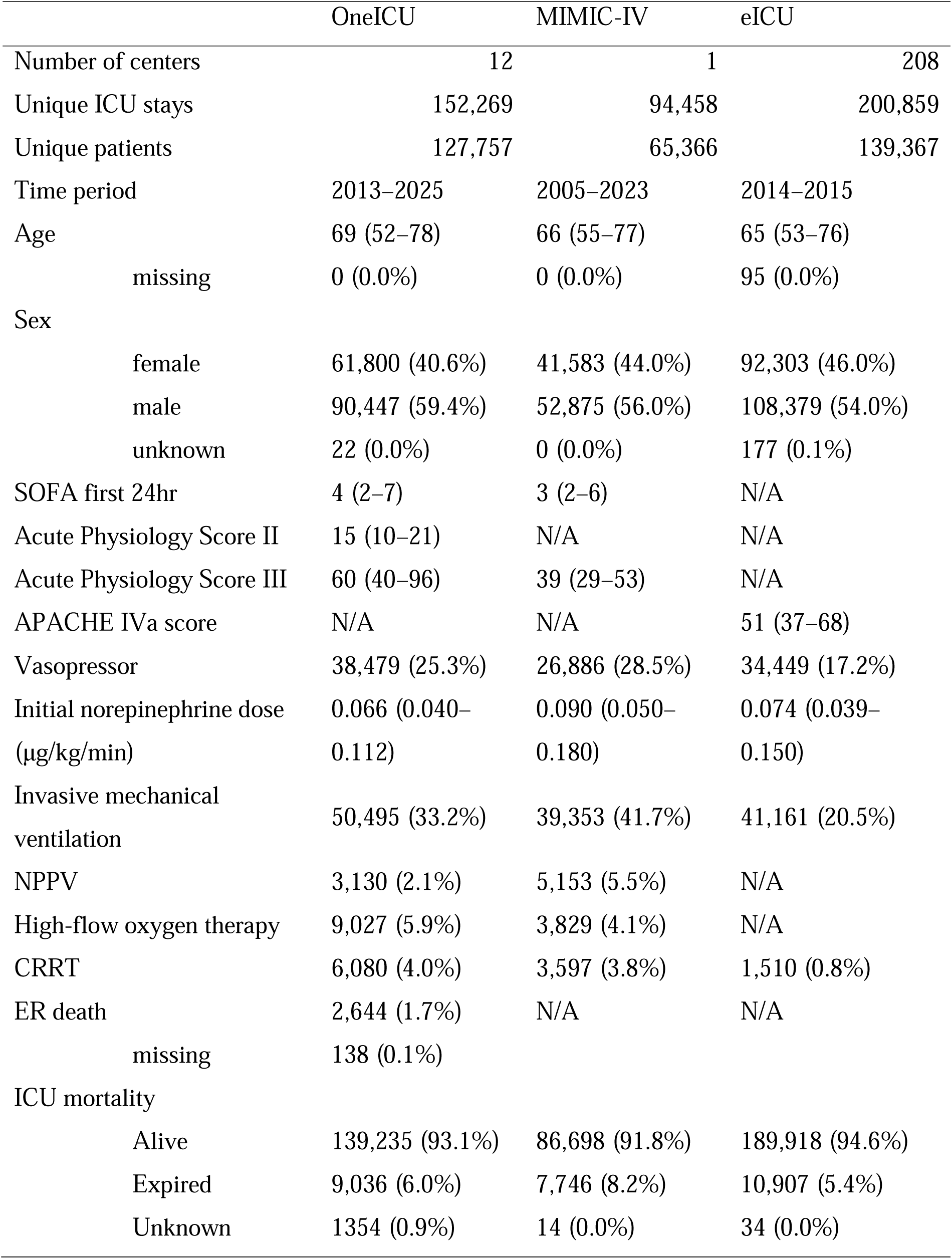
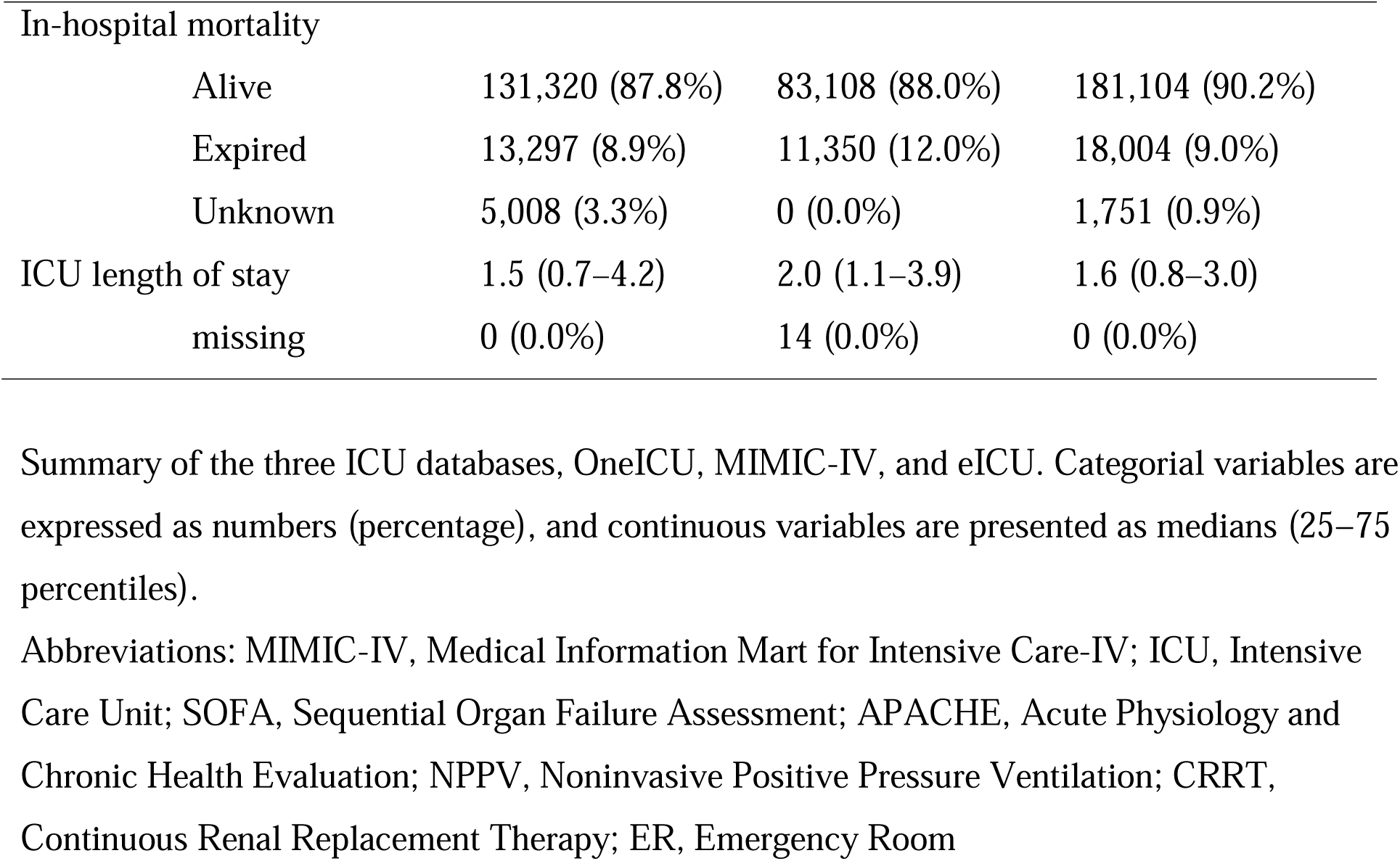
Baseline characteristics and clinical features of ICU stays in OneICU, MIMIC IV, and eICU.

|  | OneICU | MIMIC-IV | eICU |
| --- | --- | --- | --- |
| Number of centers | 12 | 1 | 208 |
| Unique ICU stays | 152,269 | 94,458 | 200,859 |
| Unique patients | 127,757 | 65,366 | 139,367 |
| Time period | 2013–2025 | 2005–2023 | 2014–2015 |
| Age | 69 (52–78) | 66 (55–77) | 65 (53–76) |
| missing | 0 (0.0%) | 0 (0.0%) | 95 (0.0%) |
| Sex |  |  |  |
| female | 61,800 (40.6%) | 41,583 (44.0%) | 92,303 (46.0%) |
| male | 90,447 (59.4%) | 52,875 (56.0%) | 108,379 (54.0%) |
| unknown | 22 (0.0%) | 0 (0.0%) | 177 (0.1%) |
| SOFA first 24hr | 4 (2–7) | 3 (2–6) | N/A |
| Acute Physiology Score II | 15 (10–21) | N/A | N/A |
| Acute Physiology Score III | 60 (40–96) | 39 (29–53) | N/A |
| APACHE IVa score | N/A | N/A | 51 (37–68) |
| Vasopressor | 38,479 (25.3%) | 26,886 (28.5%) | 34,449 (17.2%) |
| Initial norepinephrine dose<br>($\mu\text{g/kg/min}$) | 0.066 (0.040–<br>0.112) | 0.090 (0.050–<br>0.180) | 0.074 (0.039–<br>0.150) |
| Invasive mechanical<br>ventilation | 50,495 (33.2%) | 39,353 (41.7%) | 41,161 (20.5%) |
| NPPV | 3,130 (2.1%) | 5,153 (5.5%) | N/A |
| High-flow oxygen therapy | 9,027 (5.9%) | 3,829 (4.1%) | N/A |
| CRRT | 6,080 (4.0%) | 3,597 (3.8%) | 1,510 (0.8%) |
| ER death | 2,644 (1.7%) | N/A | N/A |
| missing | 138 (0.1%) |  |  |
| ICU mortality |  |  |  |
| Alive | 139,235 (93.1%) | 86,698 (91.8%) | 189,918 (94.6%) |
| Expired | 9,036 (6.0%) | 7,746 (8.2%) | 10,907 (5.4%) |
| Unknown | 1354 (0.9%) | 14 (0.0%) | 34 (0.0%) |

|  |  |  |  |  |
| --- | --- | --- | --- | --- |
| In-hospital mortality |  |  |  |  |
|  | Alive | 131,320 (87.8%) | 83,108 (88.0%) | 181,104 (90.2%) |
|  | Expired | 13,297 (8.9%) | 11,350 (12.0%) | 18,004 (9.0%) |
|  | Unknown | 5,008 (3.3%) | 0 (0.0%) | 1,751 (0.9%) |
| ICU length of stay |  | 1.5 (0.7–4.2) | 2.0 (1.1–3.9) | 1.6 (0.8–3.0) |
|  | missing | 0 (0.0%) | 14 (0.0%) | 0 (0.0%) |

Summary of the three ICU databases, OneICU, MIMIC-IV, and eICU. Categorical variables are expressed as numbers (percentage), and continuous variables are presented as medians (25–75 percentiles).
Abbreviations: MIMIC-IV, Medical Information Mart for Intensive Care-IV; ICU, Intensive Care Unit; SOFA, Sequential Organ Failure Assessment; APACHE, Acute Physiology and Chronic Health Evaluation; NPPV, Noninvasive Positive Pressure Ventilation; CRRT, Continuous Renal Replacement Therapy; ER, Emergency Room

Vital signs were measured most frequently in OneICU, followed by eICU and then MIMIC-IV (Figure 3A). Blood gas measurements (pH, lactate) were also obtained more frequently in OneICU than in the other databases, whereas complete blood count measurements (WBC, Hb) were relatively similar (Figure 3B). Albumin was measured most frequently in OneICU, while coagulation tests were performed less frequently in eICU than the other two databases.

**Figure 3.**
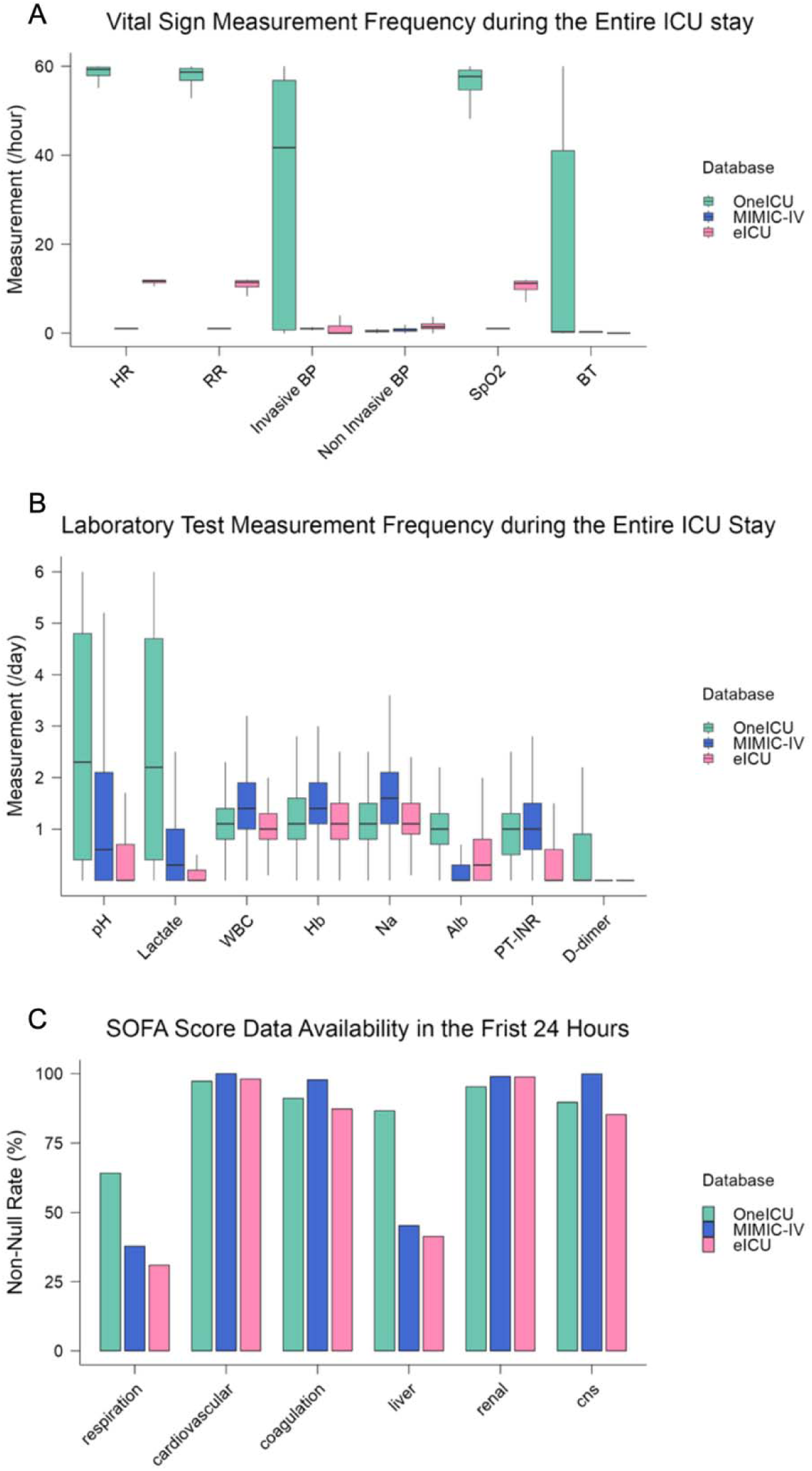
**Comparison of Measurement Frequencies Across OneICU, MIMIC-IV, and eICU** (A) Distribution of measurement frequencies for vital signs, including heart rate, respiratory rate, invasive and non-invasive blood pressure, oxygen saturation, and body temperature. OneICU records these parameters with minute-level granularity, whereas MIMIC-IV and eICU have lower measurement frequencies. (B) Comparison of laboratory test frequencies, including blood gas (pH, lactate), complete blood count (WBC, Hb), chemistry (sodium, albumin), and coagulation markers (PT-INR, D-dimer). OneICU captures these variables more frequently than MIMIC-IV and eICU. (C) Availability of SOFA score components across databases. The respiratory and liver SOFA components are measured more frequently in OneICU compared to the other databases. Abbreviations: ICU, Intensive Care Unit; MIMIC-IV, Medical Information Mart for Intensive Care-IV; HR, Heart Rate; RR, Respiratory Rate; BP, Blood Pressure; SpO , Oxygen Saturation; BT, Body Temperature; WBC, White Blood Cell count; Hb, Hemoglobin; Na, Sodium; Alb, Albumin; PT-INR, Prothrombin Time International Normalized Ratio; SOFA, Sequential Organ Failure Assessment; CNS, Central Nervous System

Cardiovascular, coagulation, renal, and central nervous system components of the SOFA score were available in over 90% of patients in all three databases (Figure 3C). Respiratory (OneICU: 73.6%; MIMIC-IV: 37.8%; eICU: 30.9%) and liver (OneICU: 93.3%; MIMIC-IV: 45.2%; eICU: 41.3%) components were captured more comprehensively in OneICU than in MIMIC-IV or eICU.

For the machine learning analysis, eligible hourly windows were approximately 2.8 million in OneICU, 4.0 million in MIMIC IV, and 1.1 million in eICU, representing 75,337, 75,668, and 32,471 unique patients, respectively. Patients were randomly assigned at the ICU-stay level to training and test sets: 60,269 (training) and 15,068 (test) in OneICU; 60,534 (training) and 15,134 (test) in MIMIC IV; and 25,976 (training) and 6,495 (test) in eICU.

Across all three datasets, AutoML selected stacked ensembles as the best performing models based on the highest internal validation AUROC. Figure 4 displays ROC curves for logistic regression and the best performing machine learning model in each dataset’s test set. The 1 minute interval model developed in OneICU achieved the highest AUROC for both approaches; the best performing machine learning model in OneICU achieved an AUROC of 0.942, the highest among all models evaluated.

**Figure 4.**
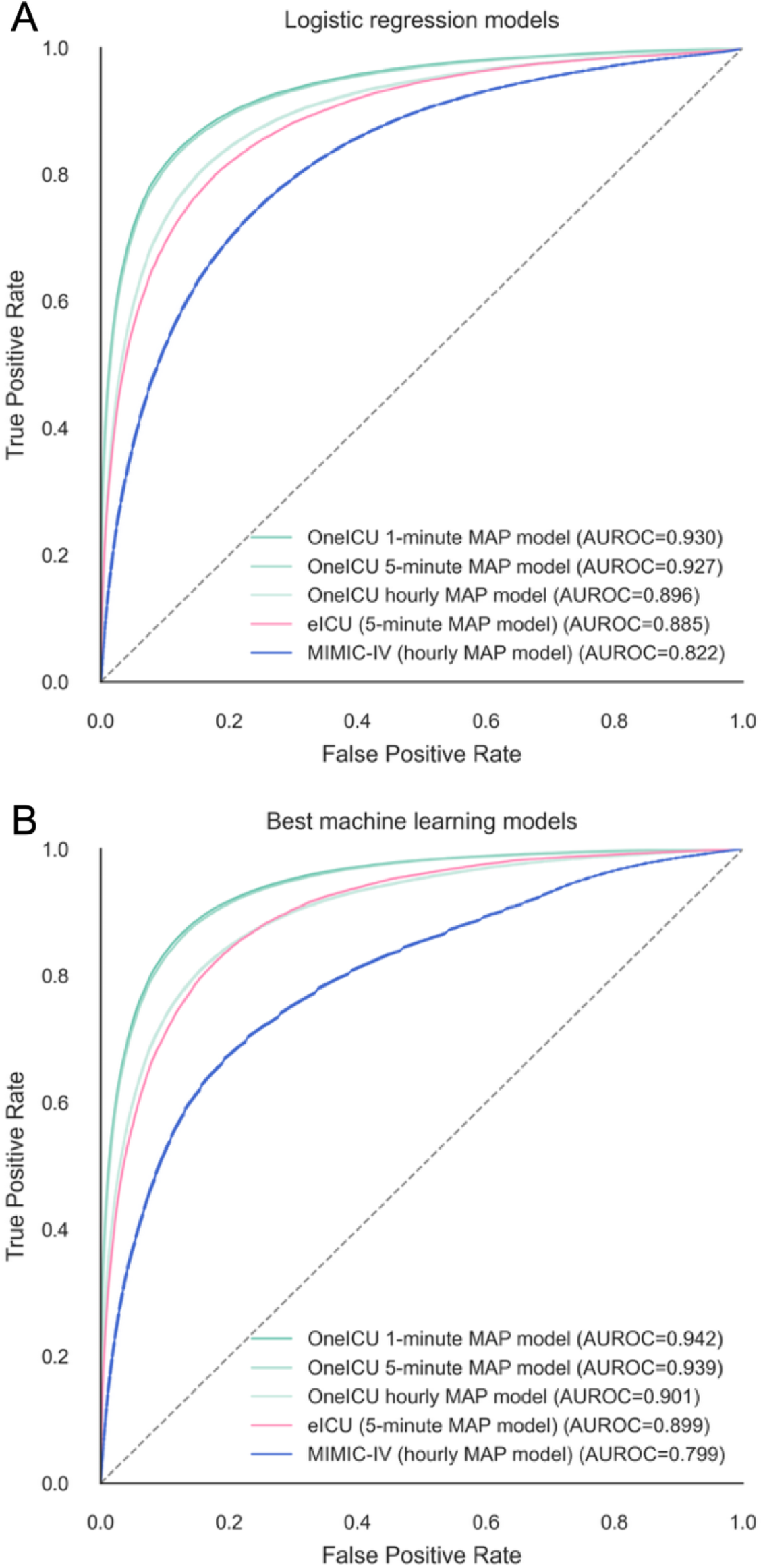
**Comparison of hypotension prediction model performance across OneICU MAP sampling frequencies, MIMIC IV, and eICU** (A) Logistic regression models using the common predictor set and database specific invasive MAP sequences (OneICU: 1 minute, 5 minute, and hourly; MIMIC IV: approximately hourly; eICU: 5 minute) (B) Best AutoML models (top ranked by AUROC on an internal leaderboard; stacked ensembles in all models) using the same predictor sets and MAP sequences as in (A) The outcome was a hypotensive event in the 60–120 minute prediction window after the index 60 minute window (median invasive MAP < 65 mmHg or vasopressor initiation). Data were split at the ICU-stay level into 80% training and 20% test sets; analytic steps were identical across databases and OneICU frequency settings except for MAP sampling frequency. Abbreviations: ICU, Intensive Care Unit; MIMIC-IV, Medical Information Mart for Intensive Care-IV; ROC, receiver operating characteristic; AUROC, area under the ROC curve; MAP, mean arterial pressure

The prevalence of hypotensive events was 12.8% in OneICU, 12.0% in MIMIC IV, and 11.3% in eICU. When sensitivity was fixed at 90%, specificity was highest for the OneICU 1 minute interval model, followed by the OneICU 5 minute interval model, the eICU 5 minute interval model, the OneICU hourly interval model, and the MIMIC IV hourly interval model (Table 2). When specificity was fixed at 90%, sensitivity was highest for the OneICU 1 minute interval model, followed by the OneICU 5 minute interval model, the OneICU hourly interval model, the eICU 5 minute interval model, and the MIMIC IV hourly interval model (Table 3). Among the three OneICU models, PPV and NPV were higher for models trained with more granular MAP measurements; comparisons of PPV and NPV across databases were not appropriate because these metrics depend on outcome prevalence.

**Table 2.**
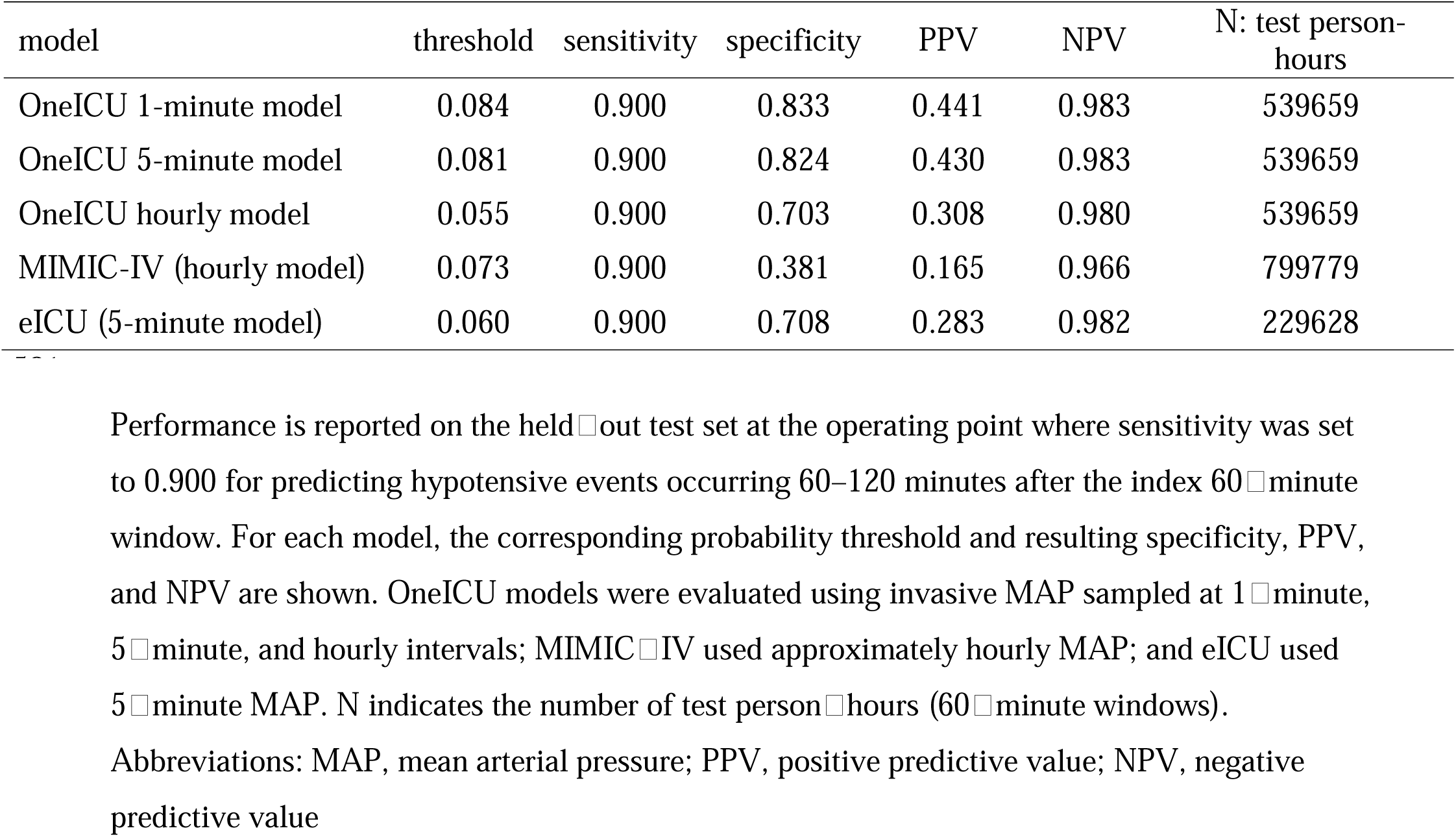
Threshold-based performance at 90% sensitivity for hypotension prediction models.

| model | threshold | sensitivity | specificity | PPV | NPV | N: test person-hours |
| --- | --- | --- | --- | --- | --- | --- |
| OneICU 1-minute model | 0.084 | 0.900 | 0.833 | 0.441 | 0.983 | 539659 |
| OneICU 5-minute model | 0.081 | 0.900 | 0.824 | 0.430 | 0.983 | 539659 |
| OneICU hourly model | 0.055 | 0.900 | 0.703 | 0.308 | 0.980 | 539659 |
| MIMIC-IV (hourly model) | 0.073 | 0.900 | 0.381 | 0.165 | 0.966 | 799779 |
| eICU (5-minute model) | 0.060 | 0.900 | 0.708 | 0.283 | 0.982 | 229628 |

Performance is reported on the held-out test set at the operating point where sensitivity was set to 0.900 for predicting hypotensive events occurring 60–120 minutes after the index 60-minute window. For each model, the corresponding probability threshold and resulting specificity, PPV, and NPV are shown. OneICU models were evaluated using invasive MAP sampled at 1-minute, 5-minute, and hourly intervals; MIMIC-IV used approximately hourly MAP; and eICU used 5-minute MAP. N indicates the number of test person-hours (60-minute windows). Abbreviations: MAP, mean arterial pressure; PPV, positive predictive value; NPV, negative predictive value

**Table 3.**
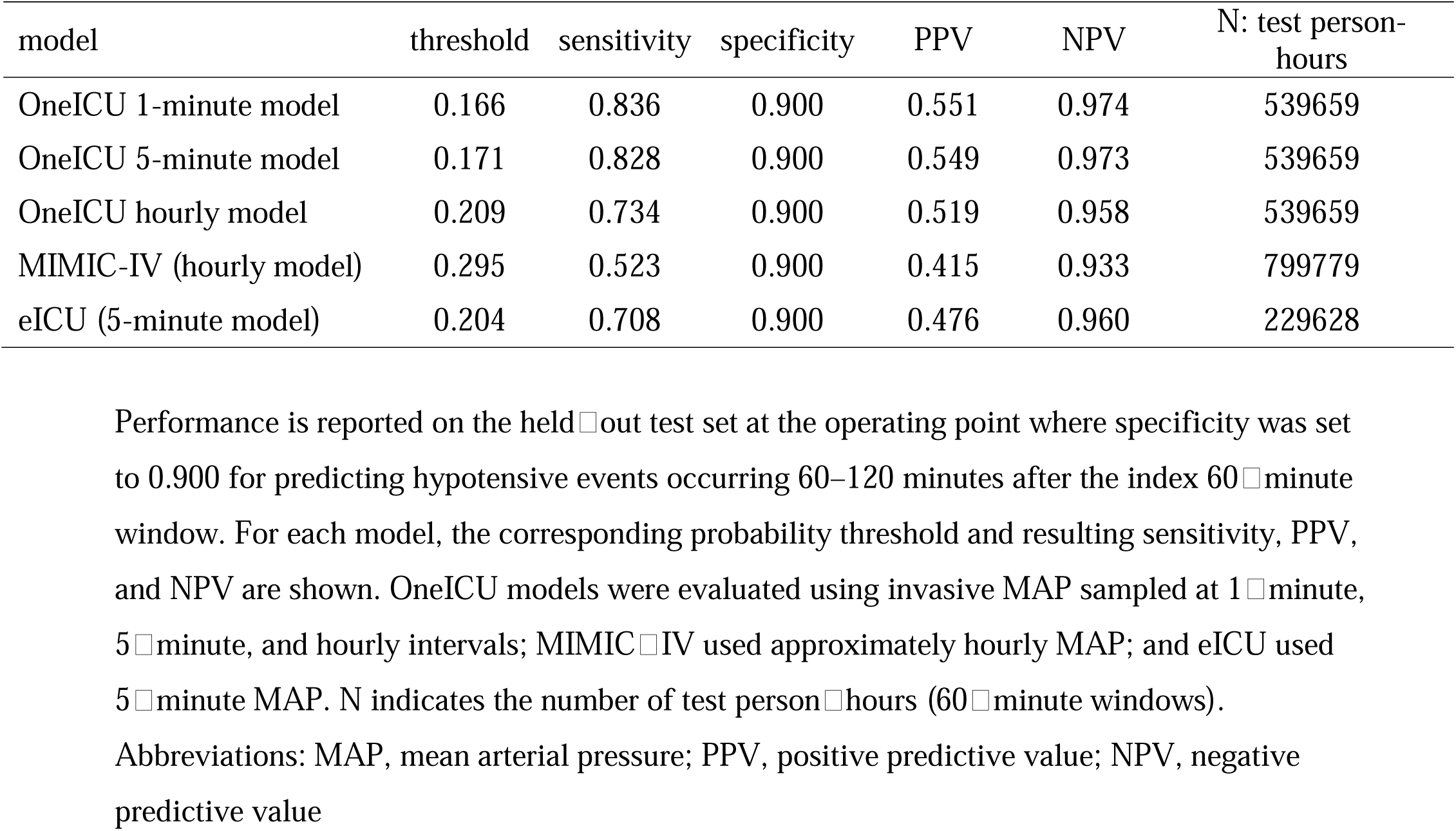
Threshold-based performance at 90% specificity for hypotension prediction models.

## Discussion

The current study describes the design and early findings of OneICU, a multicenter database that leverages minute-level ICU data captured from heterogeneous EMR systems. The high-resolution data enables the precise reconstruction of ICU clinical practice, offering insights into how patients respond to treatments and how interventions evolve over time. Figure 3 indicates that OneICU has the record of the vital signs (e.g., HR, RR, and SpO_2_) every minute in most patients, whereas the typical measurement frequency is every five minutes in eICU and every hour in MIMIC-IV. The high-frequency acquisition of core parameters allows detailed analyses of clinical trajectories, which is particularly relevant for AI-based predictive modeling.

A well-known barrier to effectively using health information technologies is the lack of EMR interoperability across multiple institutions (10,11). Differences in system architectures, data formats, and terminologies can impede the efficient data exchange among hospitals. EMR-based research databases face similar challenges due to variations in documentation principles and medical practices across institutions (12,13). For example, a study of the eICU database identified 8,565 intubated patients in the treatment table; however, records of endotracheal tube insertion and removal were available for only 1,004 of them (14). This issue likely contributes to the more frequent use of single-institution datasets such as MIMIC-III and IV, even though eICU encompasses a larger number of ICU stays overall (15).

One of the key strengths of the OneICU database is its ability to integrate diverse datasets from multiple institutions and standardize them through a streamlined ELT pipeline. By loading unstructured data onto a secure cloud-based SQL server before transformation, OneICU effectively manages variations in EMR formats, table structures, and documentation practices. This approach preserves the granular, time-stamped nature of each record while iterative transformations and standardization procedures ensure consistency across static, point time-series, and interval time-series variables. The result is a flexible, scalable schema that can accommodate continuous data growth from additional institutions, regardless of EMR vendor or hospital network, without compromising data integrity or interoperability.

We also observed that OneICU maintains a low rate of missing data for vital signs and laboratory tests despite collecting data from multiple institutions. Notably, the respiratory and liver components of the SOFA score are more commonly available in OneICU; PaO_2_ and total bilirubin are more comprehensively recorded in OneICU (16). Similarly, diagnosing DIC is far more feasible using OneICU, since fibrinogen degradation products and D-dimer, either of which is necessary for the International Society on Thrombosis and Haemostasis (ISTH)-overt DIC and JAAM DIC scoring, are rarely assessed in MIMIC-IV and eICU (17). Observed differences in availability could arise from both clinical practice and extraction process. For example, the higher computability of the SOFA liver subscore suggests that total bilirubin is measured more routinely in Japanese ICUs. By contrast, the absence of D-dimer in eICU is most consistent with non extraction rather than universal non measurement.

We also reported that vital sign sampling frequency and measurement completeness were key determinants of model accuracy in hypotension prediction. We used a strictly standardized pipeline across OneICU, MIMIC IV, and eICU, with identical inclusion and exclusion criteria, a common outcome definition, and a fixed predictor set; the only planned difference was the sampling frequency of invasive MAP. Among three models developed using OneICU, higher MAP sampling frequency was associated with higher AUROC and better threshold based metrics, including (i) specificity, PPV, and NPV at the threshold chosen to achieve 90% sensitivity and (ii) sensitivity, PPV, and NPV at the threshold chosen to achieve 90% specificity. Across datasets, AUROC was highest for the OneICU 1 minute interval model, intermediate for the eICU 5 minute interval model, and lowest for the MIMIC IV hourly interval model, consistent with each dataset’s native MAP resolution. Although direct cross database comparisons are challenging, AUROC and threshold based metrics were higher for the OneICU 5 minute interval model than for the eICU 5 minute interval model, which may reflect differences in measurement completeness of other predictors, including blood gas results. Overall, our findings suggest that higher frequency MAP measurements, together with greater measurement completeness, substantially improve the performance of machine learning models, highlighting the value of high granularity data for model development and evaluation. Developing a large-scale Asian ICU database is crucial for promoting fairness and reducing algorithmic biases in critical care research. Two recently published AI guidelines, Transparent Reporting of a multivariable model for Individual Prognosis Or Diagnosis (TRIPOD)+AI and Fairness, Universality, Traceability, Usability, Robustness, and Explainability (FUTURE)-AI, emphasize the importance of representativeness and diversity in AI model development to prevent the creation or exacerbation of health inequalities (18,19). Most established ICU databases originate from Western institutions, limiting their applicability to Asian healthcare settings characterized by different patient demographics, disease prevalence, and resource allocation. Ensuring that AI models and clinical decision-support tools are trained and validated on data representative of Asian populations not only enhances their performance in these settings but also helps prevent the perpetuation of health disparities through biased models. By systematically aggregating data from multiple institutions in Japan, OneICU takes a significant step toward equitable AI development, addressing the growing demand for robust, generalizable clinical decision-support tools that benefit critically ill patients across diverse geographical and ethnic contexts.

The present study has several limitations. First, OneICU currently includes only patients from Japanese tertiary care hospitals, the majority of whom are ethnically Japanese. Consequently, the generalizability of findings derived from this dataset to other populations may be limited. Second, the absence of imaging data and continuous waveform signals restricts research on advanced diagnostic approaches and physiological parameter monitoring. Third, while OneICU’s sequential opt-out consent process and thorough data de-identification protect patient privacy, these measures also eliminate certain demographic details and potentially relevant clinical nuances. For example, patients over 100 years old and those with rare diseases are excluded to maintain confidentiality, certain subpopulations remain underrepresented. Fourth, the models developed to compare machine learning performance across datasets are not necessarily optimal for any individual dataset, as dataset-specific features and context may have been removed by the standardized pipeline. Moreover, the generalizability of the developed models remains unclear because cross dataset testing was not feasible; models trained on higher granularity inputs cannot be applied to datasets with lower sampling resolution. Collectively, these considerations underscore the need to interpret findings derived from OneICU with caution and to pursue ongoing efforts aimed at expanding and refining this multicenter database.

## Conclusions

The OneICU database offers a high-resolution data platform for examining critical care practices and outcomes, integrating minute-level data from heterogeneous EMR systems across multiple institutions via a streamlined ELT pipeline. This capability enables detailed reconstruction of ICU workflows and advances fairness by better representing Asian populations underrepresented in existing databases.

## Declarations

### Ethics approval and consent to participate

The protocol for the original study was approved by the Kansai Medical University Ethics Committee. The current study involves the secondary use of anonymized data.

## Consent for publication

The Institutional Review Board waived the requirement for informed consent due to the anonymized nature of the data.

Availability of data and material

The authors’ agreement with the OneICU project does not permit public sharing of the data used in this manuscript. However, reasonable requests for data access will be considered by the OneICU administrator upon submission of a request through the corresponding author.

## Competing interests

TK is the founder of MeDiCU, Inc., which develops and manages the database described in this study. The other authors declare no competing interests.

## Funding

No financial support was received for this study.

## Authors’ contributions

TK conceived, designed, and coordinated the study. TK wrote the initial draft of the manuscript. YU, MW, KU, YN, SK, YI, HK, YI, AK, OT, SI, HK, and YN contributed to the interpretation of data and assisted in the preparation of the manuscript and critically reviewed the manuscript. All authors have read and approved the final manuscript.

## Data Availability

The agreement with the OneICU project does not permit public sharing of the data used in this manuscript. However, reasonable requests for data access will be considered by the OneICU administrator upon submission of a request through the corresponding author.

## Acknowledgements

None.

## Conflicts of Interests and Source of Funding

Takahiro Kinoshita is the founder of MeDiCU, Inc., which develops and manages the database described in this study. The other authors declare no conflicts of interest. No financial support was received for this study.

## Notes

### Author Declarations

The protocol for the original study was approved by the Kansai Medical University Ethics Committee (reference number: 2024015).

### Summary of Updates

Sample size increased; Manuscript updated; Figure 4 updated

