## Supplemental Methods for "Standardized Multicenter Critical Care Database Integrating Minute-Level Vital Signs, Laboratory Tests, Interventions, and Outcomes: Profile of the OneICU Database"

Supplemental Digital Content for the Profile of the OneICU Database

### Supplementary Methods

#### Overview

In this section, we describes extract–load–transform (ELT) workflow that enables efficient processing of high‑volume, unstructured data from heterogeneous EMR systems.

#### Extraction

Raw data for the OneICU database were collected from either hospital-wide or ICU-specific EMR systems at the participating hospitals, where they were archived as routine critical care records. Extracted records included demographics, administrative data, diagnoses, vital signs, physical assessments, blood gas and laboratory test results, medications, infusions, transfusions, enteral nutrition administration, medical device settings and measurements, procedures, and outcomes. All ICU examinations, measurements, and interventions were recorded with minute-level timestamps.

Neither images nor waveforms—such as those from X-rays, computed tomography, endoscopic tests, electrocardiograms, invasive arterial lines, or capnography—were collected. Free-text diagnosis records, and claim diagnoses based on the 10th revision of the International Classification of Diseases (ICD-10), were included; however, other clinical notes with high re-identification risk were excluded.

All data were de-identified immediately after extraction at the principal research institute in compliance with the Japanese Act on the Protection of Personal Information. Potentially identifiable information (names, zip codes, phone numbers, addresses, and insurance details) was securely removed and the ICU admission date was generalized to the month level, with random day assignment preserving the relative intervals among time points for each patient. Medical record numbers were encrypted using site-specific passwords and hash algorithms, ensuring both de-identification and consistency for assigning unique subject ids within the database. Potential re-identification measures, including the creation of correspondence tables, were strictly prohibited. Patients over 100 years old and those with rare diseases were excluded. These protocols were devised by lawyers specializing in personal information protection, data engineers, and intensivists, and the resulting tables were manually verified to confirm removal of potentially identifiable information.

#### Loading

Following de-identification, each record type (e.g., demographics, vital measurements, blood gas results) was consolidated into a dedicated table on a cloud-based SQL server. Because the raw data could include both long-format and wide-format structures—reflecting varying documentation practices across institutions—all data were initially standardized into JSON files containing a single column named “document.” This schema-less loading approach provided flexibility regardless of the original table format. Subsequently, each table was reshaped into a single, standardized format deemed appropriate for its specific data content.

#### Transformation

We categorized data into three hierarchical levels—patient-level, hospital-level, and ICU-level—and further classified as static, point time-series, or interval time-series variables. Patient-level data are defined as patient information invariant throughout their lifetime, including unique subject id, chromosomal sex, and date of birth. Hospital-level data are defined as patient information unique to each hospital stay, including hospital admission id, hospital id, in hospital days, hospital mortality, and ICD-10 records active during the hospitalization. ICU-level data encompass all examinations, measurements, and interventions in the ICU.

Static variables were data without repeated measurements, including age, sex, diagnosis active upon discharge from the ICU, ICU mortality, and in-hospital mortality. Point time-series data were observations or interventions performed repeatedly at distinct time points, such as vital signs, laboratory test results, physical assessments, medications, intermittent intravenous injections, and medical device measurements. Interval time-series data were variables that needed both a start and end time to reflect real-world clinical practice, such as infusion, blood transfusion, enteral nutrition, and medical device settings. During the transformation process, heterogeneous raw data were standardized into a unified structured format, leveraging these classifications.

As part of the transformation process, we implemented a natural language processing algorithm to standardize free-text diagnoses. Instead of using machine‑learning models, we adopted a rule‑based approach in which a sequence of regular‑expression rules iteratively normalized, decomposed, and generalized each diagnosis phrase. This process continued until the free-text entry aligned with recognizable disease names, and over 95% of decomposed free-text diagnoses were assigned an ICD code. Intensivists manually reviewed all final outputs to ensure semantic and clinical accuracy.

We conceptually aggregated point time-series data representing similar clinical parameters. For example, any measurements reflecting a patient's heart beat per minute—whether the heart rate (HR) recorded by electrocardiography or the pulse rate captured by a pulse oximeter—were unified under a single “heart rate” variable regardless of their sources, although it could still be disaggregated for specific research purposes. While most point time-series data were managed in a wide format, blood gas and laboratory test results were retained in a long format due to the extensive range of tests performed.

Although most raw data of interval time-series variables were recorded in long-format tables, we transformed them into wide-format tables, ensuring start and end times were properly recorded. When a clinical parameter (e.g., ventilator setting, infusion rate) changed, the corresponding record was terminated and a new record initiated.

### Schema Overview

#### patients tier:

• patients: lifetime-invariant attributes (e.g., subject_id, sex, date_of_birth, date_of_death).

#### hospitals tier:

• hospitals: site-level attributes (hospital_id, type_of_hospital)

#### hospital_admissions tier:

• hospital_admissions: single-stay attributes, including length of stay and outcomes.

• hospital_admission_diagnoses: ICD-10 admission diagnoses.

#### icu_stays tier:

• icu_stays: per-ICU-stay attributes, including ICU admission/discharge time.

##### ICU Diagnoses

• icu_diagnoses: ICU-level diagnoses (source free‑text with rule‑based mapping to ICD‑10; primary flag available).

##### Physiogical Measurements

• vital_measurements: minute-level vitals (BT, HR, RR, invasive BP, non-invasive BP, SpO_2_).

• advanced_hemodynamic_monitoring: advanced hemodynamics (cardiac output/index, pulmonary artery pressure, etc.).

• out: outputs (urine, blood loss, drains, stool, etc.).

• body_weight_measurements: body weight.

• pupillary_assessments: pupillary light reflex assessments.

##### Therapeutic Interventions

• infusions, infusion components: IV infusions with start/end times, rates, and component composition.

• injections, injection components: IV injections (time, amount, and rate) and component composition.

• blood_transfusions, blood transfusion components: blood products (including albumin) with start/end times, rates, and product details.

• enteral_nutritions, enteral nutrition components: enteral nutrition with start/end times, rates, and product composition.

• prescriptions: oral medication components.

• mechanical_ventilations: ventilator settings common to control and support modes (mode, FiO₂, PEEP) with start/end times.

• control_ventilations: controlled‑ventilation settings (inspiratory pressure, tidal volume, etc.).

• support _ventilations: support‑ventilation settings (pressure support above PEEP, percent minute volume, etc.).

• non_invasive_positive_pressure_ventilations: NIPPV settings (mode, FiO₂, PEEP) with start/end times.

• control_ventilations_nippv: controlled‑ventilation settings during NIPPV (inspiratory pressure, tidal volume, etc.).

• support_ventilations_nippv: support‑ventilation settings during NIPPV (pressure support above PEEP).

• hight_flow_oxygen_therapy: Nasal high‑flow therapy (FiO₂, total flow) with start/end times.

• oxygen_therapy: oxygen therapy (flow rate) with start/end times.

• ecmo: ECMO parameters (type, FiO₂, pump speed, sweep gas flow, blood flow) with start/end times.

• iabp: intra‑aortic balloon pump use (start/end times, assist ratio).

• impella.

• renal_replacement_therapy: RRT modality and flows.

• device_exposures: use of respiratory devices (e.g., endotracheal or tracheostomy tubes) with start/end times.

• care_unit_transfers: transfers between acute-care units (ICU, HCU, PICU, ER) with timing.

##### Laboratory & Clinical Assessments

• gcs: Glasgow Coma Scale components.

• jcs: Japan Coma Scale components.

• camicu: CAM-ICU results.

• rass: Richmond Agitation–Sedation Scale scores.

• laboratory_tests_blood: blood lab results in long format.

• laboratory_tests_others: non-blood lab results in long format (urine, cerebrospinal fluid, etc.).

• blood_gas: blood gas results in long format (ph, pO2, pCO2, lactate, etc.).

• capillary_blood_sugar: point-of-care capillary glucose.

• icu_events: ICU events such as seizures or cardiopulmonary resuscitation.

### Data Dictionary: Tables and Variables

##### patients

• subject_id — INTEGER; unique patient identifier

• female — INTEGER; biological sex (0=male, 1=female)

• date_of_birth — DATE; shifted so the first ICU admission aligns to a reference calendar

• date_of_death — STRING; shifted date of death or survival flag

##### hospitals

• hospital_ id — INTEGER; facility identifier

• type_of_hospital — STRING; public or university

##### hospital_admissions

• hospital_admission_id — INTEGER; unique hospital admission identifier

• subject_id — INTEGER; link to patients

• hospital_id — INTEGER; link to hospitals

• in_hospital_days — INTEGER; length of hospital stay [days]

• hospital_mortality — BOOLEAN; hospital mortality (true/false)

##### hospital_admission_diagnoses

• hospital_admission_id — INTEGER; link to hospital_admissions

• disease_start_date — TIMESTAMP; diagnosis start time (masked)

• disease_end_date — TIMESTAMP; diagnosis end time (masked)

• icd10 — STRING; ICD-10 diagnosis code

• suspicion_flag — BOOLEAN; suspected diagnosis indicator (true/false)

• primary — BOOLEAN; primary diagnosis (true/false)

##### icu_stays

• icu_stay_id — INTEGER; unique ICU stay identifier

• hospital_admission_id — INTEGER; link to hospital_admissions

• icu_admission_year — INTEGER; actual year of ICU admission

• icu_admission_type — STRING; emergency or scheduled_surgery

• in_time — TIMESTAMP; ICU admission time (masked)

• out_time — TIMESTAMP; ICU discharge time (masked)

• height — FLOAT; height [cm]

• er_mortality — BOOLEAN; ER mortality (true/false)

• icu_mortality — BOOLEAN; ICU mortality (true/false)

##### icu_diagnoses

• icu_stay_id — INTEGER; link to icu_stays

• diagnosis — STRING; free-text diagnosis stored in ICU system

• icd10 — STRING; mapped ICD-10 code

• primary — BOOLEAN; primary diagnosis (true/false)

##### vital_measurements

• icu_stay_id — INTEGER; link to icu_stays

• time — TIMESTAMP; timestamp (masked)

• bt_core — FLOAT; core body temperature [°C]

• bt_surface — FLOAT; surface body temperature [°C]

• hr — FLOAT; heart rate [bpm]

• rr — FLOAT; respiratory rate [breaths/min]

• non_invasive_sbp — FLOAT; cuff systolic BP [mmHg]

• non_invasive_dbp — FLOAT; cuff diastolic BP [mmHg]

• non_invasive_mbp — FLOAT; cuff mean BP [mmHg]

• invasive_sbp — FLOAT; arterial-line systolic BP [mmHg]

• invasive_dbp — FLOAT; arterial-line diastolic BP [mmHg]

• invasive_mbp — FLOAT; arterial-line mean BP [mmHg]

• spo2 — FLOAT; peripheral oxygen saturation [%]

##### advanced_hemodynamic_monitoring

• icu_stay_id — INTEGER; link to icu_stays

• time — TIMESTAMP; timestamp (masked)

• cvp — FLOAT; central venous pressure [mmHg]

• pap_systolic — FLOAT; systolic pulmonary artery pressure [mmHg]

• pap_diastolic — FLOAT; diastolic pulmonary artery pressure [mmHg]

• pap_mean — FLOAT; mean pulmonary artery pressure [mmHg]

• pawp — FLOAT; pulmonary artery wedge pressure [mmHg]

• cardiac_output — FLOAT; cardiac output [L/min]

• cardiac_index — FLOAT; cardiac index [L/min/m²]

• stroke_volume — FLOAT; stroke volume [mL]

• stroke_volume_variation — FLOAT; stroke volume variation [%]

• stroke_volume_index — FLOAT; stroke volume index [mL/m²]

• systemic_vascular_resistance — FLOAT; systemic vascular resistance [dynes⋅sec⋅cm⁻⁵]

• systemic_vascular_resistance_index — FLOAT; systemic vascular resistance index [dynes⋅sec⋅cm⁻⁵⋅m²]

##### out

• icu_stay_id — INTEGER; link to icu_stays

• time — TIMESTAMP; timestamp (masked)

• blood — FLOAT; blood loss [mL]

• urine — FLOAT; urine output [mL]

• fluid_removal_during_dialysis — FLOAT; fluid removed during dialysis [mL]

• pleural — FLOAT; pleural fluid drained [mL]

• ascites — FLOAT; ascitic fluid drained [mL]

• gastrointestinal_fluid — FLOAT; GI fluids [mL] (gastric, intestinal, bile, pancreatic)

• pericardial_fluid — FLOAT; pericardial fluid drained [mL]

• cerebrospinal_fluid — FLOAT; CSF drained [mL]

• other_body_fluid — FLOAT; other body fluids [mL]

• stool_ml — FLOAT; liquid stool [mL] (e.g., stoma output)

• stool_g — FLOAT; stool weight [g]

• urinary_frequency — FLOAT; frequency of urination [count]

• stool_frequency — FLOAT; frequency of defecation [count]

##### body_weight_measurements

• icu_stay_id — INTEGER; link to icu_stays

• time — TIMESTAMP; timestamp (masked)

• body_weight — FLOAT; weight [kg]

##### pupillary_assessments

• icu_stay_id — INTEGER; link to icu_stays

• time — TIMESTAMP; timestamp (masked)

• reflex_right — STRING; right pupillary light reflex response (positive | negative | unmeasurable)

• reflex_left — STRING; left pupillary light reflex response (positive | negative | unmeasurable)

##### infusions

• infusion_id — INTEGER; unique infusion event identifier

• icu_stay_id — INTEGER; link to icu_stays

• start_time — TIMESTAMP; start time (imputed if missing)

• end_time — TIMESTAMP; end time (imputed if missing)

• ml_per_hour — FLOAT; rate [mL/hr]

• bolus — BOOLEAN; indicator for rapid bolus

##### infusion_components

• infusion_id — INTEGER; link to infusions

• injection_product_name — STRING; product name

• unit_injection_product_per_ml — FLOAT; amount per mL

##### injections

• injection_id — INTEGER; unique injection event identifier

• icu_stay_id — INTEGER; link to icu_stays

• time — TIMESTAMP; administration time

• amount — FLOAT; volume [mL]

• ml_per_hour — FLOAT; rate [mL/hr]; may be null for slow administration

##### injection_components

• injection_id — INTEGER; link to injections

• injection_product_name — STRING; product name

• unit_injection_product_per_ml — FLOAT; amount per mL

##### blood_transfusions

• blood_transfusion_id — INTEGER; unique transfusion event identifier

• icu_stay_id — INTEGER; link to icu_stays

• start_time — TIMESTAMP; start time

• end_time — TIMESTAMP; end time

• ml_per_hour — FLOAT; rate [mL/hr]

• bolus — BOOLEAN; indicator for rapid bolus

##### blood_transfusion_components

• blood_transfusion_id — INTEGER; link to blood_transfusions

• blood_product_name — STRING; product name

• unit_product_per_ml — INTEGER; amount per mL

##### enteral_nutritions

• enteral_nutrition_id — INTEGER; unique enteral nutrition event identifier

• icu_stay_id — INTEGER; link to icu_stays

• start_time — TIMESTAMP; start time

• end_time — TIMESTAMP; end time

• ml_per_hour — FLOAT; rate [mL/hr]

• bolus — BOOLEAN; indicator for bolus administration

##### enteral_nutrition_components

• enteral_nutrition_id — INTEGER; link to enteral_nutritions

• enteral_nutrition_product_name — STRING; product name

• unit_product_per_ml — FLOAT; amount per mL

##### prescriptions

• icu_stay_id — INTEGER; link to icu_stays

• time — TIMESTAMP; timestamp (masked)

• prescription_product_name — STRING; medication product name

• amount — FLOAT; prescribed amount (units determined by product reference)

##### mechanical_ventilations

• mechanical_ventilation_id — INTEGER; unique ventilation event identifier

• icu_stay_id — INTEGER; link to icu_stays

• start_time — TIMESTAMP; start time

• end_time — TIMESTAMP; end time

• mode — STRING; ventilation mode

• fio2_ordered — FLOAT; ordered FiO2 [%] (19–100)

• peep — FLOAT; PEEP [cmH2O]

##### control_ventilations

• mechanical_ventilation_id — INTEGER; link to mechanical_ventilations

• inspiratory_pressure — FLOAT; [cmH2O]

• tidal_volume — FLOAT; [mL]

• inspiratory_time — FLOAT; seconds

• i_e_ratio — FLOAT; I:E ratio

• respiratory_rate — FLOAT; [breaths/min]

##### support_ventilations

• mechanical_ventilation_id — INTEGER; link to mechanical_ventilations

• pressure_support — FLOAT; cmH2O above PEEP

• percent_minute_volume — FLOAT; [%]

• nava_level — FLOAT; [cmH2O/μV]

##### non_invasive_positive_pressure_ventilations

• nippv_id — INTEGER; unique NIPPV event identifier

• icu_stay_id — INTEGER; link to icu_stays

• start_time — TIMESTAMP; start time

• end_time — TIMESTAMP; end time

• mode — STRING; ventilation mode

• fio2_nippv — FLOAT; FiO2 [%] (20–100)

• peep_nippv — FLOAT; [cmH2O]

##### control_ventilations_nippv

• nippv_id — INTEGER; link to non_invasive_positive_pressure_ventilations

• inspiratory_pressure_nippv — FLOAT; [cmH2O]

• tidal_volume_nippv — FLOAT; [mL]

• inspiratory_time_nippv — FLOAT; [seconds]

• respiratory_rate_nippv — FLOAT; [breaths/min]

##### support_ventilations_nippv

• nippv_id — INTEGER; link to non_invasive_positive_pressure_ventilations

• pressure_support_nippv — FLOAT; above PEEP [cmH2O]

##### hight_flow_oxygen_therapy

• icu_stay_id — INTEGER; link to icu_stays

• start_time — TIMESTAMP; start time

• end_time — TIMESTAMP; end time

• fio2_nhf — FLOAT; FiO2 [%] (20–100)

• flow_rate_nhf — FLOAT; [L/min] (1–80)

##### oxygen_therapy

• icu_stay_id — INTEGER; link to icu_stays

• start_time — TIMESTAMP; start time

• end_time — TIMESTAMP; end time

• oxygen_flow_rate — FLOAT; [L/min] (0–15)

##### ecmo

• icu_stay_id — INTEGER; link to icu_stays

• start_time — TIMESTAMP; start time

• end_time — TIMESTAMP; end time

• type — STRING; ECMO type

• fio2_ecmo — FLOAT; FiO2 [%] (20–100)

• pump_speed — FLOAT; [rpm]

• sweep_gas_flow_rate — FLOAT; [L/min]

• blood_flow_rate — FLOAT; [L/min]

##### iabp

• icu_stay_id — INTEGER; link to icu_stays

• start_time — TIMESTAMP; start time

• end_time — TIMESTAMP; end time

• assist_ratio — INTEGER; assist ratio

##### impella

• icu_stay_id — INTEGER; link to icu_stays

start_time — TIMESTAMP; start time (masked)

end_time — TIMESTAMP; end time (masked)

type — STRING; IMPELLA device type (2.5 | 5.0 | 5.5 | CP)

support_level — STRING; mechanical circulatory support level

##### renal_replacement_therapy

• icu_stay_id — INTEGER; link to icu_stays

• start_time — TIMESTAMP; start time

• end_time — TIMESTAMP; end time

• type — STRING; RRT type

• blood_flow_rate — FLOAT; [mL/min]

• body_fluid_removal_rate — FLOAT; [mL/hr]

• filtration_flow_rate — FLOAT; [mL/hr]

• dialysis_flow_rate — FLOAT; [mL/hr]

• substitution_flow_rate — FLOAT; [mL/hr]

##### device_exposures

• icu_stay_id — INTEGER; link to icu_stays

• start_time — TIMESTAMP; time when device was attached (masked)

• end_time — TIMESTAMP; time when device was detached (masked)

• device_type — STRING; type of device used

##### care_unit_transfers

• icu_stay_id — INTEGER; link to icu_stays

• time — TIMESTAMP; care unit admission time (masked)

• care_unit — STRING; care unit name (er | icu | picu | hcu | nicu | or)

##### gcs

• icu_stay_id — INTEGER; link to icu_stays

• time — TIMESTAMP; timestamp (masked)

• gcs_e — INTEGER; eye opening (1–4, C→1)

• gcs_v — INTEGER; verbal (1–5, T→1)

• gcs_m — INTEGER; motor (1–6)

• eye_opening_c — BOOLEAN; eye opening limited by swelling

• verbal_response_t — BOOLEAN; verbal response limited by tracheostomy

##### jcs

• icu_stay_id — INTEGER; link to icu_stays

• time — TIMESTAMP; timestamp (masked)

• jcs_score — INTEGER; Japan Coma Scale score (0, 1, 2, 3, 10, 20, 30, 100, 200, 300)

##### camicu

• icu_stay_id — INTEGER; link to icu_stays

• time — TIMESTAMP; timestamp (masked)

• camicu_score — STRING; (positive | negative | unmeasurable)

##### rass

• icu_stay_id — INTEGER; link to icu_stays

• time — TIMESTAMP; timestamp (masked)

• rass_score — INTEGER; (−5 to +4)

##### laboratory_tests_blood (long format)

• icu_stay_id — INTEGER; link to icu_stays

• time — TIMESTAMP; timestamp (masked)

• field_name — STRING; analyte name

• value — FLOAT; numeric value (unique per icu_stay_id, time, field_name)

• unit — STRING; reporting unit (e.g., mg/dL)

• exceeded_range — BOOLEAN; true if device exceeded measurable range

Appendix A — Common blood analytes (examples)

Hematology: rbc, hemoglobin, hematocrit, wbc, platelet.

Chemistry: total_protein, albumin, bilirubin (total/direct), AST, ALT, ALP, γ-GTP, LAP, cholinesterase, LDH, amylase, CK, CK-MB, troponin I/T, creatinine, BUN, sodium, potassium, chloride, calcium, phosphate, magnesium, uric_acid, CRP, procalcitonin, IL-6, glucose, triglyceride, total_cholesterol, HDL, LDL, BNP, NT-proBNP, ammonia, myoglobin, ferritin, HbA1c.

Coagulation: PT, PT-INR, APTT, D-dimer, FDP, fibrinogen, antithrombin.

##### laboratory_tests_others (long format)

• icu_stay_id — INTEGER; link to icu_stays

• time — TIMESTAMP; timestamp (masked)

• field_name — STRING; test item name

• value — FLOAT; numeric value (unique per icu_stay_id, time, field_name)

• unit — STRING; reporting unit (mg/dL, g/dL, meq/L, etc.)

• sample_name — STRING; specimen type (urine, cerebrospinal_fluid, pleural, ascites)

• exceeded_range — BOOLEAN; true if device exceeded measurable range

##### blood_gas (long format)

• icu_stay_id — INTEGER; link to icu_stays

• time — TIMESTAMP; timestamp (masked)

• field_name — STRING; analyte name (ph, po2, pco2, base_excess, Hb, SaO2, etc.)

• value — FLOAT; numeric value (unique per icu_stay_id, time, field_name)

• unit — STRING; reporting unit

• sample_type — STRING; sample type (arterial_blood_gas, venous_blood_gas, urine_gas, etc.)

• has_error — BOOLEAN; true if device flagged possible error

• is_adjusted — BOOLEAN; true if value adjusted

Appendix B — Common blood-gas analytes (examples)

ph, po2 (mmHg), pco2 (mmHg), base_excess (mmol/L), hemoglobin (g/dL), oxygen_saturation (%), carboxyhemoglobin (%), deoxyhemoglobin (%), oxyhemoglobin (%), methemoglobin (%), hemoglobin_f (%), total_bilirubin (mg/dL), hematocrit (%), sodium (mEq/L), potassium (mEq/L), chloride (mEq/L), calcium (mmol/L), bicarbonate (mmol/L), anion_gap (mEq/L), glucose (mg/dL), lactate (mmol/L), creatinine (mg/dL).

##### capillary_blood_sugar

• icu_stay_id — INTEGER; link to icu_stays

• time — TIMESTAMP; timestamp (masked)

• glucose_capillary — FLOAT; capillary blood glucose [mg/dL]

##### icu_events

• icu_stay_id — INTEGER; link to icu_stays

• time — TIMESTAMP; event occurrence time (masked)

• event — STRING; event type (cardiopulmonary_resuscitation | resuscitative_thoracotomy | seizure)
